# Qualitative Study of Acceptability, Benefits, and Feasibility of a Food-based Intervention among Participants and Stakeholders of the RATIONS Trial

**DOI:** 10.1101/2025.01.10.25320312

**Authors:** Sunita Sheel Bandewar, Madhavi Bhargava, Hema Pisal, Sharanya Sreekumar, Anant Bhan, Ajay Meher, Anurag Bhargava

## Abstract

**Background:** A qualitative study was conducted during the Reducing Activation of Tuberculosis by Improvement of Nutritional Status (RATIONS) trial to explore the perceptions, experiences, and expectations of participants and stakeholders on the acceptability, benefits, and feasibility of the nutritional intervention.

**Methodology:** We recruited 58 individuals for in-depth interviews (IDI) and focus group discussions (FGDs) using referential sampling. These included patients with TB (PwTB), household contacts (HHCs), and stakeholders such as trial team members, government frontline workers (the Sahiyas), and National TB Elimination Program (NTEP) staff. All IDI and FGDs were audio-recorded, transcribed, and translated. The codes were generated using an inductive process and categorized manually into themes. Direct quotes were employed to describe the themes.

**Results:** The intervention was found to be acceptable in terms of cultural compatibility, quality, quantity, and duration; considered beneficial in helping tolerate the adverse effects of medications, gain of weight, improvement of health, and resuming work; and was considered life-saving by many during the COVID-19 pandemic. Other observations included food-sharing in the control arm, inability to regain pre-disease functional status despite weight gain, and preference for either in-kind support or preference for both cash and food support in-kind. The Sahiyas favored this food-based intervention because they observed reduced TB deaths, improved adherence, and better functional recovery. They expressed confidence in its feasibility and willingness to take responsibility for its implementation. The field staff found it challenging in the context of a trial due to the added responsibilities of rigorous data collection but found it important and gratifying. The NTEP staff considered it feasible, provided the necessary resources were provided. Many favored the extension of food support to the HHCs of PwTB.

**Conclusions:** The qualitative inquiry revealed that the food-based intervention for the PwTB and their HHCs was acceptable, feasible, and beneficial for the recipients and the NTEP. Participants suggested the extension of nutritional support in some areas, as well as enhanced cash support and nutritional support to patients with all forms of TB and their HHCs. Opinion on cash or support in kind was divided; many preferred food support over cash, but others expressed a requirement for both.

## INTRODUCTION

> “The right to food is the right to have regular, permanent, and unrestricted access,. To quantitatively and qualitatively adequate and sufficient food corresponding to the cultural traditions of the people to which the consumer belongs, and which ensures a physical and mental, individual and collective, fulfilling and dignified life free of fear.”^1^

Tuberculosis (TB) is one of the most widely prevalent infections, affecting 23% of the global population.^2^ An estimated 10.8 million people fell ill with TB in 2023, with 1.25 million dying from TB in 2023, the highest with any single pathogen.^3^ As a quintessential social disease, poverty is a major factor that affects both the exposure to M. tuberculosis and the outcomes of TB infection.^4^ Undernutrition is a significant factor associated with poverty that mediates its risk of active TB, leading to immunological dysfunction, referred to as nutritionally acquired immunodeficiency syndrome.^5^ It is the leading risk factor for TB incidence worldwide, accounting for as many cases annually as due to HIV and diabetes combined.^3^

Also, undernutrition is a widely prevalent comorbidity in persons with TB (PwTB) and is a consistent risk factor for TB mortality.^6^ However, efforts in addressing this, unlike some other immunosuppressive conditions like HIV infection, are lagging. A Cochrane review was inconclusive about the effect of nutritional supplementation on treatment outcomes in PwTB.^7^ However, the WHO recommended nutritional assessment and counseling for all PwTB and support to some groups as integral components of TB care.^8^ The WHO noted a lack of evidence on undernutrition as a risk factor in household contacts (HHCs) of PwTB.^8^

The Reducing Activation of Tuberculosis by Improvement of Nutritional Status (RATIONS) trial (henceforth ‘trial’), a cluster-randomized trial of nutritional intervention in PwTB and their HHCs, in a setting with a high prevalence of poverty, food insecurity, undernutrition, and a low prevalence of HIV and drug-resistant TB, addressed this evidence gap.^9^ This was the first trial globally to estimate the effect of nutritional supplementation on TB incidence in a group at high risk of developing TB.^9^ The patient cohort in the trial was one of the largest cohorts of PwTB, which was provided nutritional support and where the clinical and nutritional outcomes were documented.^10^ This paper reports the qualitative sub-study to assess participants’ and stakeholders’ perceptions, experiences, and expectations on the acceptability and feasibility of this food basket-based intervention in the RATIONS trial.

## METHODOLOGY

### Study design

This qualitative inquiry was done at the end of the nutritional intervention period of the trial. In brief, the trial population consisted of 10,345 HHCs of 2800 adult PwTB with microbiologically confirmed TB.^11^ For ethical reasons, the documented high prevalence of severe undernutrition in PwTB in India and recommendations for nutritional care in a national policy document,^12,13,14^ patients in both trial arms received a monthly food basket and micronutrients for the treatment period (Table-1). The HHCs in the control arm (1400 families) received a food basket and micronutrients based on family size for the treatment period of the index PwTB. The primary outcome was the difference in the TB incidence in the HHCs in both arms over a two-year follow-up period.^11^

**Table 1:** RATIONS Study intervention.

|  | Intervention arm | Control arm |
| --- | --- | --- |
| Index case<br>(1200 Kcal of energy and 52 gm proteins) | 5 kg of rice<br>3 kg roasted Bengal gram powder (locally called as *sattu*)<br>1.5 kg of milk powder<br>500 ml vegetable oil<br>One RDA of micronutrient | 5 kg of rice<br>3 kg roasted Bengal gram powder (locally called as *sattu*)<br>1.5 kg of milk powder<br>500 ml vegetable oil<br>one RDA of micronutrient |
| Household contact | 5 kg rice<br>1.5kg pulses (split pigeon peas)<br>One RDA of micronutrient per adult/adolescent HHC<br>Half of this for children less than 10 years. | Nutritional counseling<br>Usual food assistance through the public distribution system |

RDA = Recommended Dietary Allowance

### Study setting and context

The trial was conducted in association with the National Tuberculosis Elimination Program (NTEP) in four districts of Jharkhand in eastern India.^11^ Jharkhand (“land of forest”) is rich in mineral resources and has a significant forest cover and a hilly terrain. According to the 2011 census, more than three-quarters of its population of 33 million lives in rural areas. As a distinctive feature of the state, more than a quarter comprises scheduled tribes (ST), a marginalized set of communities. However, two-thirds of the participants in the trial were tribals, reflecting their higher proportion in the selected districts and tuberculosis units.

Jharkhand has many tribal groups: the Mundas, Gonds, Santhals, Ho, Oraons, and Bhumij.^15^ They continue to fare the worst in terms of income, education, health, nutrition, prevalence of TB, access to health services, and nearly half are classified as suffering from Multidimensional poverty.^16^ Infant and under-five mortality, nutrition indicators like wasting and stunting in children, and thinness and anemia in adults are higher than those of the other social groups.^17^ The World Bank reported that 55% of tribal households had some degree of food insecurity throughout the year.^18^ The prevalence of TB in tribals is more than double the national average (703/100,000 vs. 316/100,000),^19,20^ In some tribal communities, like the Sahariyas, the TB prevalence is almost 10 times the national average.^21^

In Jharkhand, agriculture is the backbone of the rural economy, revolving around a single rain-fed rice crop due to the low availability of irrigation. Food insecurity is worse in the pre-harvest months of September to November due to small land holdings and subsistence farming.^18^

### Participant types

The participants in the qualitative sub-study (Table-2) included the PwTB, their HHCs, the Sahiyas, the NTEP staff, the trial field staff that anchored the regular follow-up of the families, and the food delivery, two project consultants who supervised the field staff through field visits, checks of food stocks, preparing the monthly rations procurement order, and liaising with government program staff. Sahiyas (which means a female friend in the local language) is the local name for Accredited Social Health Activists (ASHAs), the frontline health workers associated with the National Health Mission.^22^ Among many preventive and promotive health activities that the Sahiyas conduct, the TB component includes case-finding, facilitating diagnosis and treatment, and enrolment in the scheme for direct benefit transfers (DBT). DBT is a monthly cash transfer of INR 500 for all PwTB under the Nikshay Poshan Yojana to support nutritious food during treatment.^23^

**Table 2:** Profile of Study Participants for the qualitative sub-study of the RATIONS trial.

| Participant characteristics (N = 58) |  | N (Percent) |
| --- | --- | --- |
| Age group | 18-29 | 13 (22.4) |
|  | 30-39 | 32 (55.2) |
|  | 40-49 | 4 (6.9) |
|  | 50-60 | 9 (15.5) |
| Sex | Male | 34 (58.6) |
|  | Female | 24 (41.4) |
| Education | Not adequately literate | 6 (10.4) |
|  | Primary School | 5 (8.6) |
|  | High School | 21 (36.2) |
|  | Graduate | 16 (27.6) |
|  | Post Graduate | 8 (13.8) |
|  | Not mentioned | 2 (3.4) |
| Occupational status | Unemployed | 8 (13.8) |
|  | Employed | 50 (86.2) |
| Type of participants | Patients with TB | 12 (24.1) |
|  | Household contacts* | 6 (10.3) |
|  | RATIONS field staff | 16<sup>#</sup> (25.9) |
|  | Sahiyas | 18 (31.06) |
|  | RATIONS Project consultants | 2 (3.4) |
|  | NTEP Staff | 4 (1.8) |
| Type of data collection<br>(n=26) | In-depth interview | 19 (73.1) |
|  | Focused group discussion<sup>@</sup> | 4 (15.4) |
|  | Group interview | 3 (11.5) |

\* 2 Household Contacts were also Sahiyas; <sup>#</sup>1 field staff was also a TB survivor; <sup>@</sup> 6-10
Sahiyas and 8-10 field staff present in each FGD

### Research team attributes

The qualitative sub-study was anchored by social scientists from the Forum for Medical Ethics Society (FMES) with extensive experience in qualitative research and ethics. Their anchoring of the study provided expertise as well as a third-party perspective.

The lead trial investigators had experience spread over two decades of working with marginalized tribal communities in central India and researching interactions between TB and nutrition. Their long-term engagement with the NTEP, involvement in the trial conceptualization and its conduct, and experiences with the implementation of the intervention helped the stakeholder mapping, framing the in-depth interview (IDI) and focus group discussion (FGD) guide, and interpreting the data (S1 Appendix).

Interviews were conducted by HP, MB, SSB, AB, and AM in varying combinations depending on the participants of IDI and FGD.

### Sampling and recruitment strategy

The study used a referential sampling framework with participants and stakeholders across urban/rural geography, community groups, and social class. The trial field staff helped recruit PwTB and their HHCs and Sahiyas, while the trial investigators helped recruit the field staff and project consultants. The NTEP program staff were recruited with inputs from the trial team. None of the participants refused participation.

### Data collection

Data collection was done from January to June of 2022. The overall contours of the interview guide developed were adapted suitably for the respective stakeholders. The IDIs of trial participants occurred at their residences, and all FGDs were in community halls (the Panchayat Bhawan) or district headquarters. During data collection in the field, all the prevailing infection prevention guidelines of the COVID-19 pandemic were followed. All data was collected face-to-face except through online interviews with the two project consultants and one with senior NTEP staff. All interviews were conducted in Hindi, and in some, the aid of a local interpreter was utilized. The duration was 90 to 120 minutes for IDI and up to 180 minutes for FGDs.

### Data processing and analysis

All interviews were audio-recorded, transcribed, cross-checked by at least two researchers, and translated into English. Interviews were continued till HP and SS identified data saturation. For the analysis, codes (SSB, HP, and SS) were generated using an inductive process and manually categorized by the social scientists of FMES into themes. Direct quotes were employed to describe the themes.

### Ethics

Written informed consent was obtained from all participants: the PwTB, HHCs, project staff, the Sahiyas, and the NTEP staff. Appropriate privacy and confidentiality measures were undertaken, and data were anonymized. The study approvals include ICMR-NIRT: 2018020 and Ekjut Ethics Committee for local oversight and reporting.

### Measures and techniques to enhance the quality of data

There were several virtual interactions between the trial and FMES teams for developing the interview guides. To ensure rigor and credibility, the study team members known to the trial participants were absent during the interviews. Similarly, during the FGDs with Sahiyas, the government health staff were not present, and during FGD and IDI with the field staff, the lead investigators were absent.

## RESULTS

The stakeholders’ perspectives that emerged consisted of not only those related to the stated objectives like ascertainment of acceptability, content of food basket, and its benefits in terms of weight gain and ability to work but also those perspectives that were unstated aspects of the intervention. These included humane aspects like being cared for, experience of personhood, and dignity.

The results presented are organized around the themes that emerged in the content analysis and supported by relevant quotes.

### 1. Acceptability of the food basket

We explored the participants’ views regarding the acceptability of the food basket. Leading questions were avoided as far as possible. Various aspects include the cultural compatibility of the content of the food basket, palatability, ease of cooking, and the quality compared to their usual food, as described below.

#### 1.1 Cultural compatibility

Almost all participants asserted this, along with good taste. Some mentioned that ‘sattu,” although available in the market, is unaffordable and was mainly for special occasions/festivals.

> “The ration provided was compatible with our local and cultural food habits here in our society. Patients eat this kind of food only” (Female HHC)

> *“I liked all the items. … I used to make sattu balls/laddu and eat. … Milk powder, I mixed water, and it was good; I made rice and used oil to cook vegetable curry.”* (Female PwTB)

> “I knew about sattu and (also know that it) could be made as a drink or as a paste for eating. Though I was aware (but) I had never eaten sattu in the past.” (Female PwTB).

The diverse ethnic communities have their own specific food-related cultural practices. Some participants highlighted that these diverse ethnic communities have certain similarities and differences regarding their food preferences.

> “We are Munda. We have Munda, Gopa, Kurmi, Lohar (artisans engaged with fabrication and ironwork), and Santhals in this village. Eating habits (of all these ethnic / artisan groups) are almost the same but (with) some difference.” (Male PwTB)

Several meat items were desirable inclusions in the interactions with varied preferences across ethnic communities. This, along with certain commonalities, justified the content of a desirable food basket.

> “Like some eat suvar (pig) and some don’t. But most eat meat, fish, and eggs … (Therefore,) the food basket could be the same for all. All consume eggs. Santhal, Munda, Mahato, Karmakar, and even Brahmins eat eggs. (And) meat, fish can be there, but not all eat it.” (Male PwTB)

#### 1.2 Quality of the food basket, its comparison with their regular food

All trial participants unambiguously expressed satisfaction with the quality of the food basket. Many felt it was far better than what they usually eat.

> “What they gave was good and tasty. We get sattu in the market, but the sattu they gave we won’t get in the nearby shops. What I bought from a nearby shop was not good.” (Male PwTB)

We further probed the quality by comparing food baskets with what they receive from the public distribution system (PDS), one of the largest social welfare schemes of fair-priced government shops on which many poorer families across India and those in the trial rely for their food requirements. These insights are relevant as PDS can be a good strategy for scale-up in the future. Broadly speaking, three distinctive factors emerged in this comparison: the diversity of food items, the quantity, and the quality.

> “There is much difference between the rations we receive from the PDS and what they gave. (For example), they (PDS) only give rice, and here (in the trial food basket) we get sattu, milk and … The rice is also better in this.” (Male PwTB)
>
> Furthermore, his wife mentioned that the PDS must provide additional grocery items. *“We get only rice in PDS. They should give us soybean and dal, too.” (Female HHC)*

### 2. Perceived benefits of the food basket

#### 2.1 Improvement in weight and strength

The expression of benefits and gratitude varied according to the participant’s needs, vulnerabilities, and lived experiences. An important message underscored was feeling better, gaining weight, and gaining strength.

> *“I was a very thin person earlier. This food was good for me as it increased, almost doubled my weight.” (Female PwTB unemployed, physical disability, from the Indigenous community).* This was also vindicated by her daughter, who expressed a similar sentiment.
>
> “My mother became better soon with ration. We did not find it difficult to give her the food she needed (due to the food basket). She was very weak, but after getting food, her weight increased.” (Female HHC)

The benefits went beyond ‘feeling better’ to positively impact other spheres of life, such as schooling, sports, and a sense of well-being.

> “I had stopped my studies and playing on the ground when I had the disease. Now, I have started to play volleyball.” (Young male PwTB)

#### 2.2 Better adherence to treatment due to lesser side effects

Participants drew a link between “powerful” TB medicines and the favorable impact of adequate good food from their lived experiences. One patient highlighted that TB medicines could have been difficult to consume without the food basket. This is important because adherence to TB treatment is vital to achieving a cure and has important programmatic implications.

> “If I had not got the ration, it would have been very difficult for me. The medicines are so powerful that I would have died.” (Male PwTB)

This was even more eloquent from a participant who had another episode of TB in the past.

> “Earlier, when I got the disease (earlier episode), I used to have chest pain and was very weak. Now, there is no such pain, and I can work.” (Male PwTB, recurrent TB)

The complex relationship of undernutrition, TB, side effects of medicines, lack of compliance, and drug resistance was explained by a senior medical officer of tribal origin who has been working in the region for >25 years:

> “Side effects of TB medicines are more in malnourished patients. Or the situation may be that after some time, he will stop eating food due to vomiting. We will know about it only when we do the follow-up, and then we will restart medicines. This irregularity would ultimately develop drug-resistant TB.”

#### 2.3 Effects on outcomes like death and recurrence of TB

The Sahiyas, who are uniquely placed to compare patients within and outside the trial, shared their observations regarding the benefits of the food basket.

> “Those who received food baskets improved completely. They gained weight. … One of my patients was taking medicine. He migrated somewhere, but he died there. … All those who received food baskets are doing well now. But the one who did not receive it, he has got TB again.” (FGD Sahiyas)

#### 2.4 Other collateral benefits

We noted some collateral benefits of the trial team’s engagement with the families. For example, a HHC developed TB and was not eligible for a food basket. However, due to the knowledge gained about the type of diet that would be beneficial, the family attempted to maintain a diet like the trial diet. The positive impact on community food practices, thus, goes beyond TB.

> “We would not have come to know about all this, and we would not have this knowledge, such as how TB occurs, how it spreads, what food is needed to prevent it, and which medicine we should take for this disease.” (Male HHC)

The lived experiences of individuals, including non-trial participants, brought unique insights into how they perceive the relevance of such an intervention. One of the field staff, also a TB survivor, expressed this eloquently.

> “When I had TB, we had to go to STS (Senior Treatment Supervisor), and he would reply to our questions. But there is no situation where someone visits your house to help you understand the aspects of diet and lifestyle. This did not happen when I had TB. There would be large numbers of people who would not have gained weight, many malnourished.”(field staff and TB survivor)

The home visits by the field staff and periodic checkups were added benefits.

> “People in the village are very poor. They do not even have food to eat. So, the villagers never refused. Few urban families with higher incomes refused to take the food. But when we said we would come monthly for your checkup, and they agreed.” (FGD field staff)

### 3. Expectations regarding the food basket: quantity and content

> During the interaction with patients, their families, and Sahiyas, various expectations regarding the content of the food basket were expressed for a possible roll-out on a large scale. *“It is good, but some patients say that it would have been better if eggs had been given,”* was expressed by a Sahiya (FGD).
>
> Some other Sahiyas mentioned during the same FGD “Potatoes, Horlicks, Soyabean.” But then immediately, someone in the group quipped, *“Not everyone is getting the rations; if we demand too much, then whatever some are getting that also they will not get.”*

One participant, with grandchildren, who does cow grazing for livelihood and depends on his son for financial needs, wanted vegetables and meat in the basket.

> *“I will take potato, all kinds of vegetables such as tomato, brinjal then dal and rice. If there is (a possibility of getting it), I would like to have meat (once) in a week.” (Male PwTB)* However, he expressed his conviction that the government would never provide meat in the food basket. *“The government will not give us this (meat) (Laughing).”*

About the quantity, most felt it was okay, especially in the intervention arm. The trial participants in the control arm appeared to share with others, especially with children.

> “The quantity of ration they give in each area should vary by region. For example, people in the village work so hard that they need more food. But in cities, they are sitting in the workplace. They drink a lot of milk and food with good nutrients, so it is a complete food, but village people do manual work and eat less nutritious food. So, the quantity of ration given to them feels less (because they require more given their physical work). I feel it this way.” (Female HHC)

### 4. Duration of support, regaining strength, but not enough to resume work

Several study participants expressed that although the food basket was helpful and they had recovered, they have not been able to resume work, often labor-intensive, primarily because they continue to feel weak.

> “I was a mason. I felt as if having gained strength but not as much as before disease. The ration provided surely helped improve my health. … From when I am all right, I have not worked. I feel weak even now and am just staying at home.” (Male PwTB)

This suggests that the food to PwTB benefitted by being responsive to their nutritional needs. The benefits have transcended TB recovery to improve health. Yet, it was sometimes not sufficient to regain health to the extent that they could consider resuming their earlier work. Considering the high prevalence of severe undernutrition in PwTB in the trial, a significant proportion remained underweight at the end of treatment. The interventions were extended in duration in these patients.^10^ TB is more common in men, and PwTB are often the only earning members of the family, and such lean periods adversely affect the families.

> A patient with TB and diabetes, who was underweight and regained normal weight, put it across very well, *“if a person becomes healthy, then they can stop giving ration, but if someone is still weak, then they should continue to give them the food for their health.”(Male PwTB)*

A reflection of the understanding of the complexity of such an intervention by the government, the extent to which it can be done, and the general understanding of resource constraints of public spending was evident in the expression that they do not wish to receive the support longer than the health of the patient warrants.

> “It is given for the welfare of the patient so when the patient becomes healthy, it can be stopped.” (Female HHC)

### 5. Resource-constrained circumstances of households affected by TB

We encountered trial participants who never had milk in their diet. Often, participants had diet styles restricted to limited food items or only one or two meals during the day and nothing else.

> “I used to eat very little. There was not much (available with us to eat). I ate only one or two meals and used to eat very little then. … (Before receiving the food basket,) my meal used to be Alu ka sabji (potato side-dish), rice (only) this much (showing with hand).” (Male PwTB)

> “… we would not pay attention to our diet as we go to work and sometimes sleep without food only.” (Female PwTB)

Several trial participants rarely included sattu and milk in their diet. Some mentioned reasons like limited purchasing capacity, while others did not mention any specific reasons, such as an unstated dignified silence that was hard to escape the interviewers.

Limited financial resources, dependence on contractual work, often involving heavy manual labor, and overall subsistence-based livelihoods implied precarious living conditions that can easily worsen with TB. Loss of livelihood and poor strength to resume work with no reserve to sustain during the difficult times made the situation worse for these families. This became especially evident when an incident TB case developed in a household and the intervention could not be provided to them (and the DBT delayed).

> “In my house, there was very little food, and ration was also very little. Our didi (Sahiya) and everyone said you must eat good food because the medicine is very strong. Since I had no food, I became very weak…….They said I would not get a ration but would get money. That also I did not get till now.” (Male HHC)

### 6. Food basket, food insecurity and the COVID-19 pandemic

The pandemic, with the loss of livelihood and lockdown, worsened the already precarious conditions for the community and shaped the views on the intervention for many.

> “We would find it very difficult. We had to give food to my father on time, and we had also to feed everyone in the family. So when we got food, it was lifesaving.” (Female HHC)

> “I like to say that during Covid time, when we went to peoples’ homes, they used to say that since we are getting these foods, we are surviving; otherwise, we will starve here. In the trial, since we are giving rations to people, we could see sensitive issues such as hunger and poverty very closely”(Field staff).

### 7. Feasibility and mainstreaming: stakeholders’ perspective

We spoke with the participants about possible feasibility issues, and most felt that it was essential, and despite constraints, there was a willingness to contribute to the efforts.

#### 7.1 Sahiyas

They were more than willing to get involved with the logistics if this was rolled out at scale. However, they mentioned that there should be good supportive supervision and monitoring as they do not want to be accused of pilferage.

> **“**As Anganwadi workers (India has an Integrated Child Development Services Scheme where preschool children have informal education and supplementary nutrition, among other things) bring their goods, we will also unite to do the same. We are 4 in a village and will come together and do it in a tempo (small goods carrier vehicle). We will do anything for the patients.”

#### 7.2 Field staff of the trial

Apart from food delivery, the connection of the field staff with their assigned 80-120 households was an important aspect of care. This takes the issue from beyond feasibility to humane care for patients with problems like TB, where patients often experience stigma and discrimination.

> “One patient, when I took his arm to take BP, he started crying. His mother and everyone in the house started crying. When I asked him about it, he said that his doctor had never touched him in the hospital. He made me sit far away and talk and prescribed medicines to me from a distance. You two are the first people who have touched me and are talking to me face to face.”

The field staff did express difficulty distributing the food basket on top of rigorous data collection of a research trial. During the peak of the trial, with enrolment and follow-up of already enrolled patients, the difficult-to-reach areas or the scattered hamlets were especially challenging. But gratification was also expressed by most.

> “The geographical situation here is such that the villages are scattered. So how many people can you visit in one day? You cannot visit 8-10 patient’s homes in 14 days.” (field staff FGD).

> “When you do something for someone, and you see the positive things happening because of you, it feels good. Every job provides you money, but only a few provide ultimate pride and satisfaction.” (field staff FGD)

For feasibility, it is important to mention the acceptability of project field staff delivering food baskets at the doorstep. While most participants were very happy to have it delivered, there were challenges initially with some families. Villagers would gather and ask many questions due to trust issues. Things improved gradually with the improvement in the health of the trial participants, appropriate explanations by Sahiyas, and other sociocultural methods employed by the field staff.

> “We would speak their language, drink their water, eat food that they offered, and then they would accept us. They start trusting us once we help them with an illness and facilitate their investigations.”(field staff FGD)

#### 7.3 NTEP staff

The feasibility aspect was also discussed with STS, especially if the intervention was scaled up and included all PwTB at the district level.

> He said, *“Even if done on a big scale, I do not think it should cause many problems because patient numbers will slowly reduce (with this intervention).”*

Another senior NTEP staff felt this is very much needed but that the challenges he foresees are procurement, storage, and distribution, which can be overcome by budgeting appropriately for human resources.

### 8. Who should receive? The equity lens

The equity aspect of who should get it was discussed in various ways: microbiologically confirmed PwTB, all PwTB, and HHCs.

Sahiyas expressed that it was uncomfortable for them as they were dealing with patients who were part of the trial as well as outside the trial:

> “All patients should get food. If food is given to all, due to the availability of nutritious food and medicines, the patient will recover quickly, and the disease will not spread. Most of them are poor, so all should get.”(Sahiya FGD)

> “I think food should also be given to the patient’s family. Because even if the patient is cured, the weak family member will get TB. We do not want to spread TB; we want to control it. Food is important for (preventing) this, so give it in advance.”

> “In my village, one TB patient told me that you are giving food to that patient. Is he your friend? And I am your enemy, so you are doing this. We hear this kind of thing.”

The sense of community in rural areas is higher compared to urban areas, and this was expressed by field staff.

> “People in villages mostly do farming, so there is a lot of exchange going on among them. If one has less, 10 people help him. They have more love, affection, and tighter bonds.”

Food sharing was especially common in the control arm, with a female index patient, when there were toddlers or more children in the family.

> One of the grandmothers, who was also a patient, said, *“I give the milk powder to my grandson when I have it” (Female PwTB)*

For extending food to HHCs, the responses were “yes” and “no,” each qualified appropriately.

> “No, the nutritious food is for the patients and their health. So, why should it be given to the whole family? Patients are taking the medicine, and there are side effects. So, they should take nutritious food.” (Female HHC).

On further probing these divergent opinions, one trial participant provided a response that conveyed her qualified choices and preferences rooted in the understanding of economic constraints. The resource constraints and possible non-feasibility for the government in supporting the food provision for the entire household of all PwTB might explain this divergence in the response. This was despite their own microcosms against the backdrop of difficult personal circumstances and limited financial capabilities.

### 9. Cash and kind

Most participants felt that both cash and food in kind are important. But when asked to choose, many felt that food is better than cash which may get spent on things other than nutrition.

> “Considering the present situation, Rs 500/-is not much for the patients. Maybe this amount needs to be increased at least by Rs 200/-more.” (STS)

> “I would choose food and medicine. To get well, we need medicine, and to become healthy, we need food. I don’t need money if I get the food and medicines…….it will be difficult for TB patients to go outside and buy those (food items) needed for their health. It would be helpful if someone gave them food on time” (Male PwTB)

The divergent opinion was also based on the presumption that patients will use the money on unnecessary expenses, including the possibility of alcohol. But at the same time, cash could be used for the food of their choice.

> “If we had gotten money, then we could buy chicken and all.” (Male PwTB).

However, it is important to note that branded and chocolate-flavored powders advertised as energy boosters were commonly mentioned during many interviews.

### 10. Stigma

Stigma can be reflected in keeping distance from TB patients and their families, and at times, it could even lead to ostracism – subtle or obvious. Stigma was more prominent in urban areas than in rural and tribal communities.

Unsurprisingly, we found misconceptions about TB, at least amongst certain sections. Insights from one of the field staff, also a TB survivor, were helpful.

> “A girl living nearby had attained the age of marriage. Their whole family, including the girl, did not disclose her disease status as it would hamper her prospects of marriage.

> They told us not to visit their house with ration and other items and they came and collected it.” (Female field staff, TB survivor)

However, we also heard voices stating that once TB was cured, there was not much to worry about the marriage prospects of TB patients, especially in the rural and tribal areas.

> “All this (stigma) is present among educated people.” (Tribal Medical Officer)

Practices to prevent the spread of infection can also be construed as stigma or can give rise to perceived stigma by the patient.

> “A person with TB should stay separately from other people at home. The person should use different vessels for eating and drinking water, and other family members should not use the vessel he is using. He should not spit around anywhere. He should spit only in a polythene bag and put it in the garbage or should burn the polythene bag to ashes.”

We probed the possible stigma related to ration being delivered.

> “Nothing like that. Even when brother (field staff) used to come to give food, neighbors used to ask why they were coming. We used to just tell them the reason.” (Male PwTB)

## DISCUSSION

The qualitative inquiry on various aspects of the food-based intervention with participants and other stakeholders in the RATIONS trial reaffirmed the felt need for such a food-based intervention. There are valuable additional insights into the composition of the food basket, the acceptability, the feasibility, the perceived benefits, and the background conditions, especially poor nutrition and food insecurity of the participants. Many participants alluded to food insecurity, manifesting in the unaffordability of nutrient-dense foods like sattu and milk powder to even cutting back on the number of meals consistent with moderate to severe food insecurity definitions by FAO.^24^ Undernutrition is most commonly due to food insecurity, which “is the limited or uncertain availability of nutritionally adequate, safe foods or the inability to acquire foods in socially acceptable ways.”^25^ The trial had >80% of the PwTB,^10^ and >1/3^rd^ HHCs with undernutrition.^9^ The background to this is poverty, in which people develop TB, which is further exacerbated by the inability of the breadwinner to work, creating a vicious cycle.

Food insecurity is widely prevalent in India,^26^ including moderate to severe food insecurity in households with PwTB,^25,27^ which is linked with many adverse outcomes.^28^ In the recent comprehensive national nutrition survey, 3 of 5 adolescents did not have access to nutritious foods like fruit or milk even once a week.^29^ In the National Family Health Survey-5, more than half of adults did not consume any pulses or beans daily, nearly half did not consume any dairy products or dark green vegetables, more than 80% did not consume any fruits, and more than 90% did not consume any eggs or flesh foods.^30^ Household food insecurity was strongly associated with the development of active TB in children (aOR: 11.55).^27^ Food insecurity is also linked to treatment adherence,^31^ and clinical outcomes in PwTB, with a fivefold higher risk of death.^28^ The need for a food basket in such a situation was evident by the term “life-saving” used by some trial participants. Food insecurity has also been linked to a higher prevalence of mental health issues such as depression and anxiety, which has an impact on treatment success.^32^

The acceptability of the food baskets was uniformly high and attributed to content and quality. However, the perceptions about their quantity and duration varied according to individual needs and lived experiences. The composition of the trial food basket was based on the NTEP policy document.^14^ The recommendations were based on deliberations of a multi-disciplinary group of experts in nutrition science, dietetics, clinical medicine, tuberculosis, clinical researchers, and program implementation.^14^ Discussions were held with a local not-for-profit organization in Jharkhand and the community health workers in which Sattu emerged as a locally available and acceptable protein-rich food. The basket focused on meeting energy and protein needs with the recommended daily allowance of micronutrients through the micronutrient pills. Similar food baskets based on dry food rations containing cereals and pulses have been well accepted in West Bengal and Madhya Pradesh in India.^33–35^. Also, food baskets have been used with good acceptability in Afghanistan,^36^ Senegal,^37^ Angola,^38^, and Brazil.^39^

In India, under the National Food Security Act of 2013, 75% of rural households and 50% of rural households have access to cheaper rice and wheat in PDS. However, PDS meets the cereal requirements of people and assumes that the resultant savings will improve their non-cereal intake.^40^ The predominantly poor participants liked the quality and diversity of our food baskets. The basket gave them the equivalent of 500 ml of milk and 100 gm of pulses per day (Sattu). Sattu is versatile and provides choices in terms of consumption. It can be made into a savory or sweet drink, can be rolled into sweet balls, or stuffed into freshly baked savory Indian bread (Paratha). The diversity breaks the monotony that can occur with ready-to-use therapeutic foods (RUTF). Such locally available and acceptable sources of proteins need to be identified for state-level scale-up.

The responses to the perceived benefits of the food baskets included a range of outcomes relevant not just to the patients but also to the NTEP. These include becoming productive members of society, better adherence, reduced mortality, and reduced side effects. A very striking feature in Indian PwTB patients is the high prevalence and severity of undernutrition.^10,12,13,41^ Undernutrition and poor weight gain are associated with a higher risk of death, drug toxicity, and recurrence of TB even after a cure.^6,42,43^ Significantly, in the setting of severe weight loss and poor weight gain, the ability to return to work is impaired. Many patients reported an increase in strength to return to their usual activity. However, the persistence of undernutrition (40% continued to remain underweight at 6 months in the trial because of severe undernutrition at baseline),^10^ and the presence of post-TB sequelae explains why some patients reported an inability to work as usual at the end of treatment.

It was also encouraging to see Sahiyas report anecdotally a lower risk of death and relapse in trial patients as compared to those outside the trial. Food baskets served as an enabler to better adherence to drugs that were otherwise difficult to tolerate. This is apart from the general observation that food assistance is associated with improved adherence to antitubercular therapy.^44^ The role of food insecurity in increasing the frequency and intensity of adverse drug reactions (ADR) has been noted in studies in other countries, including in patients with HIV disease.^45,31^

Many patients tolerated the therapy better due to the food baskets. Undernutrition is a risk factor for drug toxicity with anti-TB drugs, but there is a paucity of ADR studies on PwTB and undernutrition. A cohort of tribal patients in a community with a high burden of TB and undernutrition documented ADR in nearly 90% of PwTB, much more than that in our trial.^46,10^ In a recent study in children and adolescents, ADRs were strongly associated with malnutrition, with Grade-3 ADRs exclusively seen in patients with malnutrition.^47^

Apart from providing the food baskets, the participants appreciated the nutritional counseling the trial field staff provided. Counseling augments the understanding of a balanced diet, addresses certain myths, and does not adversely affect the long-term eating habits in a community. There are discussions to introduce peanut-based energy-dense nutrition supplements (EDNS)/RUTF in India for PwTB. A study of the acceptability of peanut-based EDNS in 102 PwTB over a 2-month period reported a 10% refusal and nearly 14% reported adverse effects.^48^ The recommended dose of this EDNS results in more than 60gm fat and 28gm protein per day.^48^ This would not address the additional protein requirement in an illness like TB and lost lean body mass. Also, the high-fat content may be responsible for nausea and fat accretion rather than lean mass. In a small study in Africa of RUTF vs. food baskets in PwTB, nearly half reported a preference for food baskets.^37^ In studies with peanut-based RUTF in Bangladesh, 40% of caregivers of children and nearly 80% of women found it unacceptable as a food product, citing palatability and smell despite the perception of a therapeutic benefit.^49,50^ In Indian children, too, the acceptability of a ‘khichri’ (cooked preparation of rice and legumes) was much higher than that of a peanut-based RUTF.^51^ RUTFs risk mystifying nutrition and making local communities dependent on higher-priced externally sourced food items.

The responses to questions of duration and provision of food baskets reflected a nuanced understanding of the feasibility of support. There is a need for an extension of support when needed. The study also explored views regarding the composition of the food basket, and participants did mention other desirable items, like vegetables, fruits, and eggs/meat, with the understanding that they may not be feasible.

Regarding the feasibility of implementing nutritional support, there was strong support from the Sahiyas with their first-hand observation of the beneficial effects on the treatment outcomes. Their willingness was with the caveat of community involvement and supportive supervision of the process. Additionally, the Sahiyas handed over a written petition at the end of FGD to convey this wish to the NTEP. The trial field staff provided valuable insights into the challenges of making doorstep deliveries of food rations in challenging terrain and hard-to-reach areas. The additional task of trial data collection and measurements further complicated their work.

We also elicited the perspectives regarding cash transfers, an operationally easier alternative to facilitate purchasing nutritious foods. The DBT by NTEP operational since 2018 has been christened the Nikshay Poshan Yojana.^23^ The PwTB in India work in the unorganized sector and depend on daily wages, with no paid sick leave, continued wages, or comprehensive health insurance covering outpatient and inpatient care. Studies done more than two decades ago showed significant levels of indebtedness, largely due to indirect costs associated with loss of wages in Indian PwTB.^52^ In a recent national cost survey, >45% of PwTB incurred catastrophic costs (costs >20% of annual household income).^53^ The question of cash transfers vs. food support in kind drew responses that either supported in-kind food support or the provision of both food and cash. It was felt that the current provision of Rs. 500/month was inadequate to either address the financial needs or purchase a nutritious diet. Although an impact evaluation of the NPY has shown that non-receipt of DBT was associated with more than 4-fold higher odds of unfavorable outcomes,^54^ there has been no evaluation of the nutritional impact of the DBT. As a positive step, the cash benefit for PwTB is to be doubled.^55^ An important step should be assessing the nutritional impact of these initiatives to shape future policies.

Finally, the qualitative study highlighted some persistent misconceptions that can impact rational and cost-effective nutritional care: the desire to buy branded energy powders with little value for money. This is because of the aggressive marketing of such products in the print and audio-visual media and reinforces the need to add counseling as an important long-term enabler in TB-care.

### Strengths and limitations

This qualitative inquiry had numerous strengths. First, it was conducted among a population whose voices are often under-represented in the literature because they are rural, live in remote areas, and belong to indigenous communities. Second, when qualitative aspects are studied in trials, there is potential bias when studied by the trial investigators. The presence of FMES social scientists unrelated to the trial helped minimize this. Third, it was ensured that the researchers or the field staff who knew the participants were not present during the interviews/discussions. Fourth, most in-person interactions were at the homes of the trial participants which helped shape the understanding of the background conditions, the terrain, and the non-verbal communication that can be affected by online methods. Lastly, interim findings from the quantitative trial component were available to shape the interview guide.

The study had some limitations. The referential sampling may not have fully represented the spectrum of perceptions, experiences, and opinions. The interviews were conducted primarily in Hindi, and some of the nuances that may have been expressed in the local language may have been affected. This study did not explore gender issues related to nutrition and food. The interviews were conducted with adults and did not reflect the experience of children and adolescents.

## CONCLUSIONS

The qualitative sub-study of the RATIONS trial revealed that the food-based intervention for the PwTB and their HHCs was acceptable in terms of content, quantity, and duration. However, there were suggestions for extended nutritional support for some. The lived experiences expressed as weight gain, recovery of strength to work, ability to adhere to treatment because of better tolerability of drugs, and gratitude are important not just for the PwTB but also for the NTEP. The field staff felt that the doorstep delivery was feasible but was challenging when combined with the data collection for the trial. The Sahiyas, with their experience of seeing PwTB within and outside the trial, felt strongly in favor of this intervention. They were willing to contribute, provided there was community oversight. The participant responses indicated an understanding of the limitations of the government regarding quantum, beneficiaries, and duration. Opinion on cash and kind was divided; many preferred food over cash, but others expressed a requirement for both.

## Data Availability

All relevant data are within the paper and its Supporting Information files.

## ACKNOWLEDGEMENT

The authors would like to express appreciation for the tremendous efforts of the entire RATIONS field team, who worked in difficult terrain and continued work without interruption during the COVID-19 pandemic. The cooperation of the district TB officers and their teams is gratefully acknowledged.

## SUPPLEMENTARY APPENDIX

1. Guidelines and steps followed for the In-depth Interview (IDI) and Focus Group Discussion (FGD)

## REFERENCES

1. The fight for the right to food: lessons learned. Basingstoke: New York: Palgrave Macmillan; 2011.

2. Houben RM, Dodd PJ. The Global Burden of Latent Tuberculosis Infection: A Re-estimation Using Mathematical Modelling. PLoS Med. 2016 Oct 25;13(10):e1002152. doi: 10.1371/journal.pmed.1002152.

3. Global Tuberculosis Report 2024. Geneva: World Health Organization, 2024. [cited 2024 Dec 14]. Available from: https://www.who.int/teams/global-tuberculosis-programme/tb-reports/global-tuberculosis-report-2024

4. Bhargava A, Bhargava M, Juneja A. Social determinants of tuberculosis: context, framework, and the way forward to ending TB in India. Expert Rev Respir Med. 2021 Jul;15(7):867–883. doi: 10.1080/17476348.2021.1832469.

5. Bhargava A. Undernutrition, nutritionally acquired immunodeficiency, and tuberculosis control. BMJ. 2016 Oct 12;355:i5407. doi: 10.1136/bmj.i5407.

6. Waitt CJ, Squire SB. A systematic review of risk factors for death in adults during and after tuberculosis treatment. Int J Tuberc Lung Dis. 2011 Jul;15(7):871–85. doi: 10.5588/ijtld.10.0352.

7. Grobler L, Nagpal S, Sudarsanam TD, Sinclair D. Nutritional supplements for people being treated for active tuberculosis. Cochrane Database Syst Rev. 2016 Jun 29;2016(6):CD006086. doi: 10.1002/14651858.CD006086.pub4.

8. Guideline: Nutritional care and support for patients with tuberculosis. Geneva: World Health Organisation, 2013. Available from: https://www.who.int/publications/i/item/9789241506410.

9. Bhargava A, Bhargava M, Meher A, Benedetti A, Velayutham B, Sai Teja G, et al. Nutritional supplementation to prevent tuberculosis incidence in household contacts of patients with pulmonary tuberculosis in India (RATIONS): a field-based, open-label, cluster-randomised, controlled trial. Lancet. 2023 Aug 19;402(10402):627–640. doi: 10.1016/S0140-6736(23)01231-X. Epub 2023 Aug 8. Erratum in: Lancet. 2024 Jul 27;404(10450):340. doi: 10.1016/S0140-6736(24)01500-9.

10. Bhargava A, Bhargava M, Meher A, Teja GS, Velayutham B, Watson B, et al. Nutritional support for adult patients with microbiologically confirmed pulmonary tuberculosis: outcomes in a programmatic cohort nested within the RATIONS trial in Jharkhand, India. Lancet Glob Health. 2023 Sep;11(9):e1402–e1411. doi: 10.1016/S2214-109X(23)00324-8.

11. Bhargava A, Bhargava M, Velayutham B, Thiruvengadam K, Watson B, Kulkarni B, et al. The RATIONS (Reducing Activation of Tuberculosis by Improvement of Nutritional Status) study: a cluster randomised trial of nutritional support (food rations) to reduce TB incidence in household contacts of patients with microbiologically confirmed pulmonary tuberculosis in communities with a high prevalence of undernutrition, Jharkhand, India. BMJ Open. 2021 May 20;11(5):e047210. doi: 10.1136/bmjopen-2020-047210.

12. Padmapriyadarsini C, Shobana M, Lakshmi M, Beena T, Swaminathan S. Undernutrition & tuberculosis in India: Situation analysis & the way forward. Indian J Med Res. 2016 Jul;144(1):11–20. doi: 10.4103/0971-5916.193278.

13. Bhargava A, Chatterjee M, Jain Y, Chatterjee B, Kataria A, Bhargava M, Kataria R, D’Souza R, Jain R, Benedetti A, Pai M, Menzies D. Nutritional status of adult patients with pulmonary tuberculosis in rural central India and its association with mortality. PLoS One. 2013 Oct 24;8(10):e77979. doi: 10.1371/journal.pone.0077979.

14. Guidance document on nutritional care and support for patients with tuberculosis in India. Central TB Division, Ministry of Health and Family Welfare,Government of India; 2017. Available from: https://tbcindia-wp.azurewebsites.net/guidance-document-nutritional-care-support-for-tb-patients-in-india/

15. Tribes of Jharkhand. 2024/12/19/T15:20:15Z 2024. Available from: https://en.wikipedia.org/w/index.php?title=Tribes_of_Jharkhand&oldid=1263951585files/3982/Tribes_of_Jharkhand.html (accessed 28th December 2024).

16. National Multidimensional Poverty Index: Baseline report. New Delhi: Niti Aayog, 2021. Available from: https://www.niti.gov.in/sites/default/files/2021-11/National_MPI_India-11242021.pdf.

17. Tribal Health Report, India – First Comprehensive Report on Tribal Health in India. Available from: https://tribalhealthreport.in/.

18. South Asia Food and Nutrition Security Initiative. Food and nutrition security in tribal areas in India. In: World Bank SAg, editor.; 2014. Available from: https://documents1.worldbank.org/curated/en/620611468173967141/pdf/Food-and-nutrition-security-in-tribal-and-backwards-areas-in-India.pdf

19. Thomas BE, Adinarayanan S, Manogaran C, Swaminathan S. Pulmonary tuberculosis among tribals in India: A systematic review & meta-analysis. Indian J Med Res. 2015 May;141(5):614–23. doi: 10.4103/0971-5916.159545.

20. National TB Prevalence Survey in India (2019-2021): Summary Report: Indian Council of Medical Research (ICMR) New Delhi, ICMR-National Institute for Research in Tuberculosis (NIRT) Chennai, Ministry of Health and Family Welfare (MOHFW) Government of India New Delhi, Central TB Division (CTD) and National Tuberculosis Elimination Programme (NTEP), World Health Organisation(WHO) India Office New Delhi, 2022. Available from: https://tbcindia.mohfw.gov.in/2023/06/06/national-tb-prevalence-survey-in-india-2019-2021/

21. Rao VG, Bhat J, Yadav R, Muniyandi M, Sharma R, Bhondeley MK. Pulmonary tuberculosis - a health problem amongst Saharia tribe in Madhya Pradesh. Indian J Med Res. 2015 May;141(5):630–5. doi: 10.4103/0971-5916.159560.

22. National Health Mission. 2024/09/15/T05:56:15Z 2024. Available from: https://en.wikipedia.org/w/index.php?title=National_Health_Mission&oldid=1245802579files/3984/National_Health_Mission.html (accessed 28th December 2024).

23. Direct Benefit Transfer Manual for National Tuberculosis Elimination Programme. In: Central TB Division, Ministry of Health and Family Welfare, Government of India, editors. New Delhi, India; July 2020.

24. Hunger and food insecurity | FAO | Food and Agriculture Organization of the United Nations. 2024. Available from: https://www.fao.org/hunger/en (accessed 28th December 2024).

25. Ayiraveetil R, Sarkar S, Chinnakali P, Jeyashree K, Vijayageetha M, Thekkur P, et al. Household food insecurity among patients with pulmonary tuberculosis and its associated factors in South India: a cross-sectional analysis. BMJ Open. 2020 Feb 28;10(2):e033798. doi: 10.1136/bmjopen-2019-033798.

26. McKay FH, Sims A, van der Pligt P. Measuring Food Insecurity in India: A Systematic Review of the Current Evidence. Curr Nutr Rep. 2023 Jun;12(2):358–367. doi: 10.1007/s13668-023-00470-3.

27. Jubulis J, Kinikar A, Ithape M, Khandave M, Dixit S, Hotalkar S, et al. Modifiable risk factors associated with tuberculosis disease in children in Pune, India. Int J Tuberc Lung Dis. 2014 Feb;18(2):198–204. doi: 10.5588/ijtld.13.0314.

28. Richterman A, Saintilien E, St-Cyr M, Claudia Gracia L, Sauer S, Pierre I, et al. Food Insecurity at Tuberculosis Treatment Initiation Is Associated With Clinical Outcomes in Rural Haiti: A Prospective Cohort Study. Clin Infect Dis. 2024 Aug 16;79(2):534–541. doi: 10.1093/cid/ciae252.

29. Ministry of Health and Family Welfare (MoHFW), Government of India, UNICEF, Population Council. Comprehensive National Nutrition Survey (CNNS) National Report. New Delhi, 2019. Available from: https://nhm.gov.in/WriteReadData/l892s/1405796031571201348.pdf

30. Patwardhan. S, R. Kapoor, S. Scott, P.H. Nguyen, S. Chamois, S.K. Singh, et al. 2022. Trends and patterns in consumption of foods among Indian adults: Insights from National Family Health Surveys, 2005-06 to 2019-21. POSHAN Data Note 91. New Delhi, India:International Food Policy Research InstituteAvailable from: https://ebrary.ifpri.org/utils/getfile/collection/p15738coll2/id/136530/filename/136743.pdf

31. de Pee S, Grede N, Mehra D, Bloem MW. The enabling effect of food assistance in improving adherence and/or treatment completion for antiretroviral therapy and tuberculosis treatment: a literature review. AIDS Behav. 2014 Oct;18 Suppl 5:S531–41. doi: 10.1007/s10461-014-0730-2.

32. Wang Q, Dima M, Ho-Foster A, Molebatsi K, Modongo C, Zetola NM, Shin SS. The association of household food insecurity and HIV infection with common mental disorders among newly diagnosed tuberculosis patients in Botswana. Public Health Nutr. 2022 Apr;25(4):913–921. doi: 10.1017/S1368980020004139.

33. Samuel B, Volkmann T, Cornelius S, Mukhopadhay S, MejoJose, Mitra K, et al. Relationship between Nutritional Support and Tuberculosis Treatment Outcomes in West Bengal, India. J Tuberc Res. 2016 Dec;4(4):213–219. doi: 10.4236/jtr.2016.44023.

34. Singh AK, Siddhanta A, Goswami L. Improving tuberculosis treatment success rate through nutrition supplements and counselling: Findings from a pilot intervention in India. Clinical Epidemiology and Global Health 2021; 11:100782.

35. Howell E, Dammala RR, Pandey P, Strouse D, Sharma A, Rao N, et al. Evaluation of a results-based financing nutrition intervention for tuberculosis patients in Madhya Pradesh, India, implemented during the COVID-19 pandemic. BMC Glob Public Health. 2023 Sep 4;1(1):13. doi: 10.1186/s44263-023-00013-6.

36. Pedrazzoli D, Houben RM, Grede N, de Pee S, Boccia D. Food assistance to tuberculosis patients: lessons from Afghanistan. Public Health Action. 2016 Jun 21;6(2):147–53. doi: 10.5588/pha.15.0076.

37. Benzekri NA, Sambou JF, Tamba IT, Diatta JP, Sall I, Cisse O, et al. Nutrition support for HIV-TB co-infected adults in Senegal, West Africa: A randomized pilot implementation study. PLoS One. 2019 Jul 18;14(7):e0219118. doi: 10.1371/journal.pone.0219118.

38. Santos E, Felgueiras Ó, Oliveira O, Duarte R. The Effect of a Basic Basket on Tuberculosis Treatment Outcome in the Huambo Province, Angola. Arch Bronconeumol (Engl Ed). 2018 Mar;54(3):167–168. English, Spanish. doi: 10.1016/j.arbres.2017.08.013.

39. Cantalice Filho JP. Food baskets given to tuberculosis patients at a primary health care clinic in the city of Duque de Caxias, Brazil: effect on treatment outcomes. J Bras Pneumol. 2009 Oct;35(10):992–7. English, Portuguese. doi: 10.1590/s1806-37132009001000008.

40. Ghosh SM, Qadeer I. Impact of Public Distribution System on Quality and Diversity of Food Consumption. Economic and Political Weekly. 2021 Jan 30;56(5):52–9.

41. Mahapatra A, Thiruvengadam K, Nair D, Padmapriyadarsini C, Thomas B, Pati S, et al. Effectiveness of food supplement on treatment outcomes and quality of life in pulmonary tuberculosis: Phased implementation approach. PLoS One. 2024 Jul 16;19(7):e0305855. doi: 10.1371/journal.pone.0305855.

42. Yew WW, Leung CC. Antituberculosis drugs and hepatotoxicity. Respirology. 2006 Nov;11(6):699-707. doi: 10.1111/j.1440-1843.2006.00941.x.

43. Khan A, Sterling TR, Reves R, Vernon A, Horsburgh CR. Lack of weight gain and relapse risk in a large tuberculosis treatment trial. Am J Respir Crit Care Med. 2006 Aug 1;174(3):344–8. doi: 10.1164/rccm.200511-1834OC.

44. Wagnew F, Gray D, Tsheten T, Kelly M, Clements ACA, Alene KA. Effectiveness of nutritional support to improve treatment adherence in patients with tuberculosis: a systematic review. Nutr Rev. 2024 Sep 1;82(9):1216–1225. doi: 10.1093/nutrit/nuad120.

45. Weiser SD, Tuller DM, Frongillo EA, Senkungu J, Mukiibi N, Bangsberg DR. Food insecurity as a barrier to sustained antiretroviral therapy adherence in Uganda. PLoS One. 2010 Apr 28;5(4):e10340. doi: 10.1371/journal.pone.0010340.

46. Mishra P, Bhat J, Yadav R, Sharma RK, Rao VG. Adverse Drug Reaction Patterns of First-line Anti-tubercular Drugs among Saharia Tuberculosis Patients: An Observational Study in Particularly Vulnerable Tribal Group of Madhya Pradesh, India. Indian J Public Health. 2023 Oct 1;67(4):542–545. doi: 10.4103/ijph.ijph_865_22.

47. Agarwal A, H B S, Mathur SB, Kalra BS, Arora R, Khanna A, Rajeshwari K. Adverse drug reactions in children and adolescents on daily antitubercular regimen: An observational longitudinal study. Indian J Tuberc. 2023;70 Suppl 1:S76–S81. doi: 10.1016/j.ijtb.2023.07.004.

48. Kumar R, Krishnan A, Singh M, Singh UB, Singh A, Guleria R. Acceptability and Adherence to Peanut-Based Energy-Dense Nutritional Supplement Among Adult Malnourished Pulmonary Tuberculosis Patients in Ballabgarh Block of Haryana, India. Food Nutr Bull. 2020 Dec;41(4):438–445. doi: 10.1177/0379572120952306.

49. Ali E, Zachariah R, Shams Z, Manzi M, Akter T, Alders P, et al. Peanut-based ready-to-use therapeutic food: how acceptable and tolerated is it among malnourished pregnant and lactating women in Bangladesh? Matern Child Nutr. 2015 Oct;11(4):1028–35. doi: 10.1111/mcn.12050.

50. Ali E, Zachariah R, Dahmane A, Van den Boogaard W, Shams Z, Akter T, et al. Peanut-based ready-to-use therapeutic food: acceptability among malnourished children and community workers in Bangladesh. Public Health Action. 2013 Jun 21;3(2):128–35. doi: 10.5588/pha.12.0077.

51. Dube B, Rongsen T, Mazumder S, Taneja S, Rafiqui F, Bhandari N, Bhan MK. Comparison of Ready-to-Use Therapeutic Food with cereal legume-based khichri among malnourished children. Indian Pediatr. 2009 May;46(5):383–8..

52. Rajeswari R, Balasubramanian R, Muniyandi M, Geetharamani S, Thresa X, Venkatesan P. Socio-economic impact of tuberculosis on patients and family in India. Int J Tuberc Lung Dis. 1999 Oct;3(10):869–77.

53. Jeyashree K, Thangaraj JWV, Shanmugasundaram D, Giridharan SLP, Pandey S, Shanmugasundaram P, et al. Cost of TB care and equity in distribution of catastrophic TB care costs across income quintiles in India. Glob Health Res Policy. 2024 Dec 9;9(1):51. doi: 10.1186/s41256-024-00392-9.

54. Jeyashree K, Thangaraj JWV, Shanmugasundaram D, Sri Lakshmi Priya G, Pandey S, Janagaraj V, et al. Evaluation Group N. Ni-kshay Poshan Yojana: receipt and utilization among persons with TB notified under the National TB Elimination Program in India, 2022. Glob Health Action. 2024 Dec 31;17(1):2363300. doi: 10.1080/16549716.2024.2363300.

55. Union Health Minister Unveils Key Initiatives to boost Nutrition Support for TB Patients and their Families. 2024. Available from: https://pib.gov.in/pib.gov.in/Pressreleaseshare.aspx?PRID=2062928files/3993/PressReleaseIframePage.html (accessed 3rd January 2025).

